# A prospective controlled study for EEG records comparison between Scalp-EEG and Ear-EEG wearable device, for subsequent analysis by an artificial intelligence-based system: SERAS-EEG study

**DOI:** 10.1101/2023.10.01.23296029

**Authors:** G. Torres-Gaona, D. Blánquez, A. Valls, X. Raurich, J. Valls, L. Munsó, J.L. Arcos, A. Trejo, A. Aledo-Serrano

## Abstract

mjn-SERAS is an earpiece shaped as a hearing-aid device, which continuously records the electrical brain activity in two channels placed in the external auditory. It uses an artificial intelligence algorithm (AI), based system for early detection of preictal period of seizures. Sixteen patients with drug-resistant focal epilepsy and 14 control subjects were simultaneously studied with the mjn-SERAS device and a standard 24-channel EEG using the 10-20 system. Data from channels F8-T4 or F7-T3, according to the laterality of the epileptic focus was extracted from the standard EEG. We analyzed the average signal correlation (AC) between the two types of records, with and without artefact removal (filtered records [FR]), comparing inter-subject and subjects recordings (SR), as well between ictal and interictal periods in epilepsy patients.

AC was 0.90 [0.88 - 0.91] and 0.88 [0.86 - 0.90] in the FR and the whole cohort, respectively. No differences in the correlation of signals were found between controls and patients in the FR (-0.01 [-0.04;0.01], p=0.261) or the SR (-0.03 [-0.06;0.01], p=0.09). In addition, in the subset of patients with epilepsy, no differences in AC were noted between interictal activity and seizures (-0.02 [- 0.06; 0.02], p=0.352). Only AC during sleep in controls was found to be smaller compared to repose (-0.04 [-0.08;-0.01], p=0.01). No adverse events were reported. Our study supports an adequate correlation between the information recorded with both methods, providing technical support for use of the mjn-SERAS to record EEG signals.

## 1. INTRODUCTION

Electroencephalography (EEG) is the main non-invasive diagnostic method in epilepsy, allowing classification and localization, and assessing the potential risk of seizure recurrence (1). Furthermore, prolonged video-EEG (v-EEG) monitoring, by analyzing both epileptic activity and ictal seizure semiology, often allows confirmation of the diagnosis and follow-up monitoring of the epilepsy (2,3). However, this method has limited sensitivity, requires a long analysis time and is subject to various interobserver and intraobserver biases (4). Moreover, although EEG is widely used, some important EEG applications such as long-term monitoring of neurological patients, sleep monitoring and brain-computer interfaces (BCI), are limited by the accessibility and mobility requirements of the equipment used. Although subcutaneous electrodes, currently approved in Europe, provide useful and good quality information, they represent a problem for patients with apprehension about surgery and need to be removed after a period of use. In addition, a clinical feasibility study to assess a long-term implanted seizure advisory system designed to predict seizure likelihood and quantify seizures served as a starting point for this purpose (5).

Non-invasive devices, such as wristbands, chestbands, headphones, and headsets, represent convenient and user-friendly options due to their discreet and extended usage capabilities. These non-invasive, long-term monitoring solutions offer the potential for seizure registration, detection, and prediction. Numerous EEG recording methods have been proposed, some of which provide relatively unobstructed access to the ear area, making them robust, cost-effective, unobtrusive, and suitable for monitoring brain activity beyond the confines of a laboratory setting (6).

Ear-EEG, a form of encephalographic recording utilizing small devices placed in the ear canal, employs dry sensors and digitization systems to convey neural data. Ear-EEG devices yield signals closely resembling the electrical source and have been under investigation for several years (9). While these devices typically feature fewer channels than a standard 10-20 setup, they are not designed for diagnosing or localizing epileptogenic zones. Instead, they serve complementary functions like monitoring, quantification, seizure detection, and prediction. Signal quality hinges not only on the electrodes employed but also on the device’s acquisition and connection system(7)

Seizure prediction holds significant clinical and functional value for patients and their caregivers, enhancing user safety, fostering independence, and facilitating improved comprehension by healthcare professionals. This, in turn, leads to more precise treatment adjustments.

mjn-SERAS solution, resembling a hearing aid, continuously monitors brain electrical activity via three sensors. It communicates via Bluetooth and employs a software application in the user’s mobile phone with an artificial intelligence (AI)-based algorithm, developed from EEG signal analysis, to enable early detection of seizures. The device can generate an individual-specific algorithm (8). Prior studies have assessed the effectiveness of ear EEG in comparison to traditional v-EEG (9). In the SERAS-EEG study, we explore the correlation between conventional v-EEG records and mjn-SERAS to identify preictal and interictal segments in patients with drug-resistant epilepsy.

## 2. METHODS

In this prospective unblinded and controlled study aimed at comparing EEG records between v-EEG and the mjn-SERAS system, we enrolled participants (patients and healthy subjects) of both sexes aged between 12 and 65 years. The patient group included individuals with a confirmed diagnosis of drug resistant epilepsy. Exclusion criteria for the patient group were the presence of dissociative seizures, severe psychiatric, neurological or systemic disorders or more than 10 seizures per day.

From each subject, we recorded simultaneously with the mjn-SERAS system and v-EEG 21-channel recordings using the 10-20 system. Subsequently, we extracted data from channels F8-T4 or F7-T3, according to the laterality of the epileptic focus in the patients’ group, and we computed the average signal correlation (AC) between the two types of recordings.

In the patients’ group, the EEG recordings were visually analyzed by specialized clinical staff of the Neurosciences department of the Corachan Clinic Barcelona, registering the number, duration and lateralization of seizures. Artifact annotation was performed by visual inspection of the signal and the video.

We analyzed the AC in the full recordings and after removing the epochs affected visually by artifacts (filtered records). In the study, we examined the intersubject and intrasubject differences of the AC. Additionally, in the patient group we studied the AC between ictal and interictal segments, while in the control group between wakefulness at rest or performing cognitive tasks (writing, reading, calculation) and sleep.

This study was carried out in Barcelona, in accordance with the recommendations of the regional ethics committee with written informed consent from all subjects. All subjects gave written informed consent in accordance with the Declaration of Helsinki. The protocol was approved by the Regional Ethical Drug Investigational Committee of Madrid’s community, Reference: 47/916513.9.9/19

### 2.1. Video-EEG acquisition and electrode positioning

The Natus Seizure Advisory System was chosen to record video-EEG data, employing a comprehensive scalp montage. The EEG data consisted of 21 channels recorded using the 10-20 system, which employs gold-plated disc electrodes affixed with collodion adhesive. Additionally, EKG and video recordings were obtained. Electrode impedances were carefully maintained below 10 kilo-ohms at the beginning of the recording. The EEG was captured using a referential montage at a sampling rate of 500 Hz. In the group of patients, for in site seizure detection, the recorded data underwent visualization and inspection with high-pass and low-pass filters set at 0,5Hz and 35 Hz or 70 Hz, respectively. Sensitivity was adjusted between 5 and 10 μV/mm. In most recordings, a 50-Hz notch filter was applied. Trained epilepsy monitoring unit (EMU) nurses were responsible for pressing the alarm button and documenting notes regarding clinical seizures or other significant events. For the chosen channels, an approximation to the mjn-SERAS device was employed, using either F7-T3 and T3-T5 or F8-T4 and T4-T6, depending on the location of focus.

### 2.2. Video-EEG analysis pre-processing

Before analysis all EEG data was reviewed the day following the recording by epilepsy-specialized neurologists with experience as electroencephalographers (G.T.); using a Natus viewer standard visual inspection, and 20s per page.

The analysis of the EEG records was performed offline after the recordings, using *Neurowork Natus* with data filter from 0.3Hz to 50 Hz. Labelling of seizures was performed by an experienced epileptologist, using the evaluation of video and scalp-EEG.

EEG records were exported from Natus to EDF file to process time series with Python algorithms, synchronizing the timeline bases. The spectrogram functions have been performed with *Python software* by creating time-frequency-power plots from short-time moving window sources of the Fourier transform.

### 2.3 mjn-SERAS Medical Device

The mjn-SERAS device is a portable device recording the brain electrical activity using three sensors, 2 channels and 1 reference placed inside the external auditory canal. The device analyzes the data collected with a mathematical algorithm that is able to determine the possibility of presenting epileptic seizures, issuing an alert to the user at least one minute before the seizure that allows the user to take safety measures in case of such an event (Fig. 1). The device has the CE mark for Europe, by notified body BSI Group, CE 685187.

**Figure 1.**
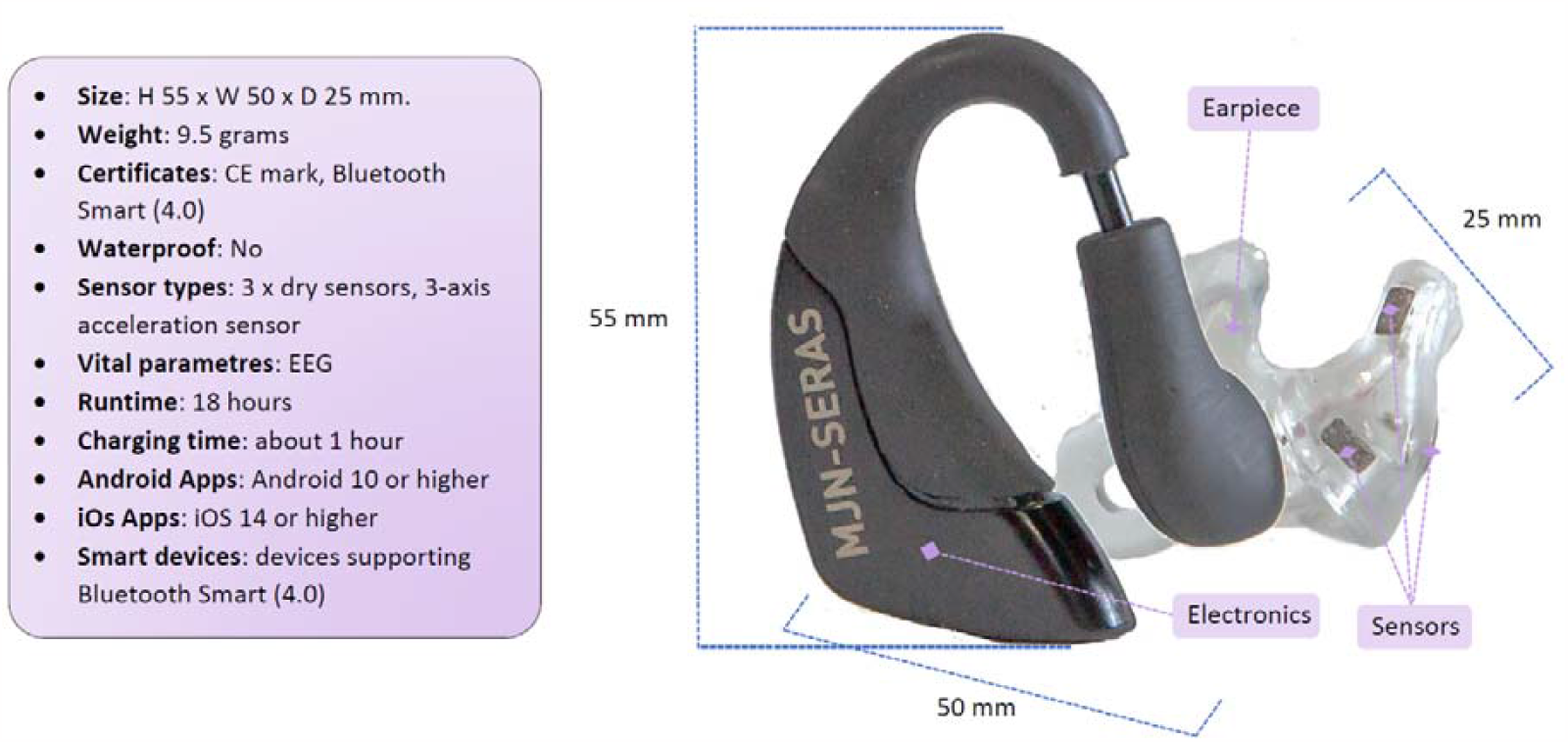
Specifications for the device

The mjn-SERAS app is designed to be used by patients with epilepsy on their own mobile phone, linked to the wearable headset. The main functionality is the seizure risk display. Once the screen goes to HIGH risk, the app displays a seizure alert message (visual, vibrating and acoustic) and the user must check that he/she has received it. For the next 15 or 30 minutes, the app will remain silent waiting for the event, and after this period it will ask the patient about the presence of the seizure, to confirm it. Users can manually add a seizure if the system is in a training period. This period depends on the seizure frequency and requires at least 4 to 5 seizures to personalize the algorithm. Clinicians can access seizure reports, seizure representations and recorded data from their patients. The device only provides monitoring information and does not have any treatment effect.

Ear-EEG was recorded with the mjn-SERAS device at a sample rate of 125Hz, useful information up to 62 Hz with a notch filter for 50Hz in European countries. Two channels were recorded and stored with a timeline base in web servers, accredited for medical data storage with anonymized identifiers. Due to the short distance between the electrodes of the ear-EEG mjn-SERAS, the amplitudes of these signals are always between 10 and 20 dB lower than those of v-EEG (13). Also, most of the information employed by algorithms is concentrated in the low frequency bands (fig 2). To compensate these signal displays, filters in the 0.3 Hz-10 Hz and 0.3Hz-4Hz bands and a 10-20 dB amplification were employed in the ear-EEG signal.

**Figure 2:**
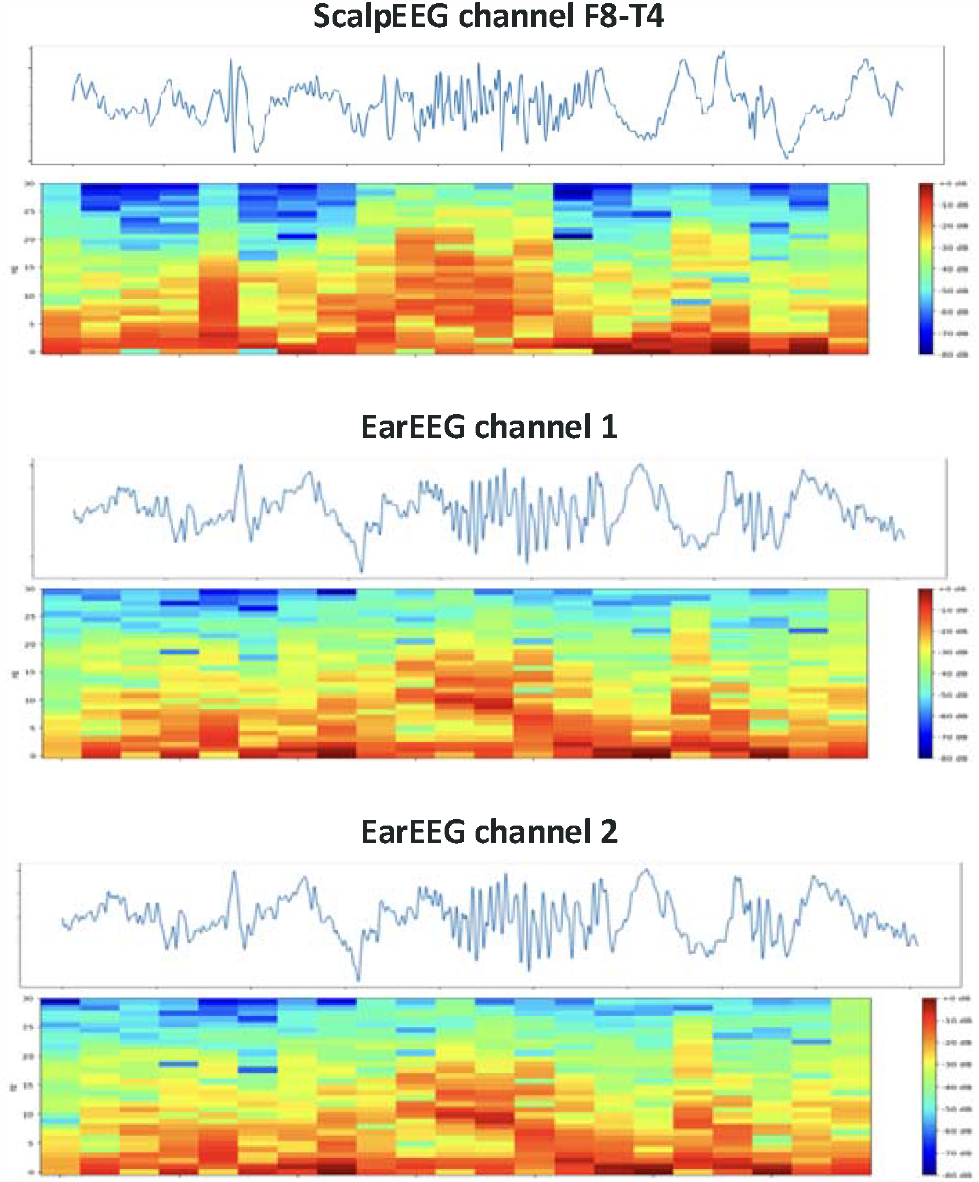
Spectrogram of the signals collected by both methods, in a 30-second section with 2-second intervals, spectrogram with blue background and red activity.

**Fig. 3.**
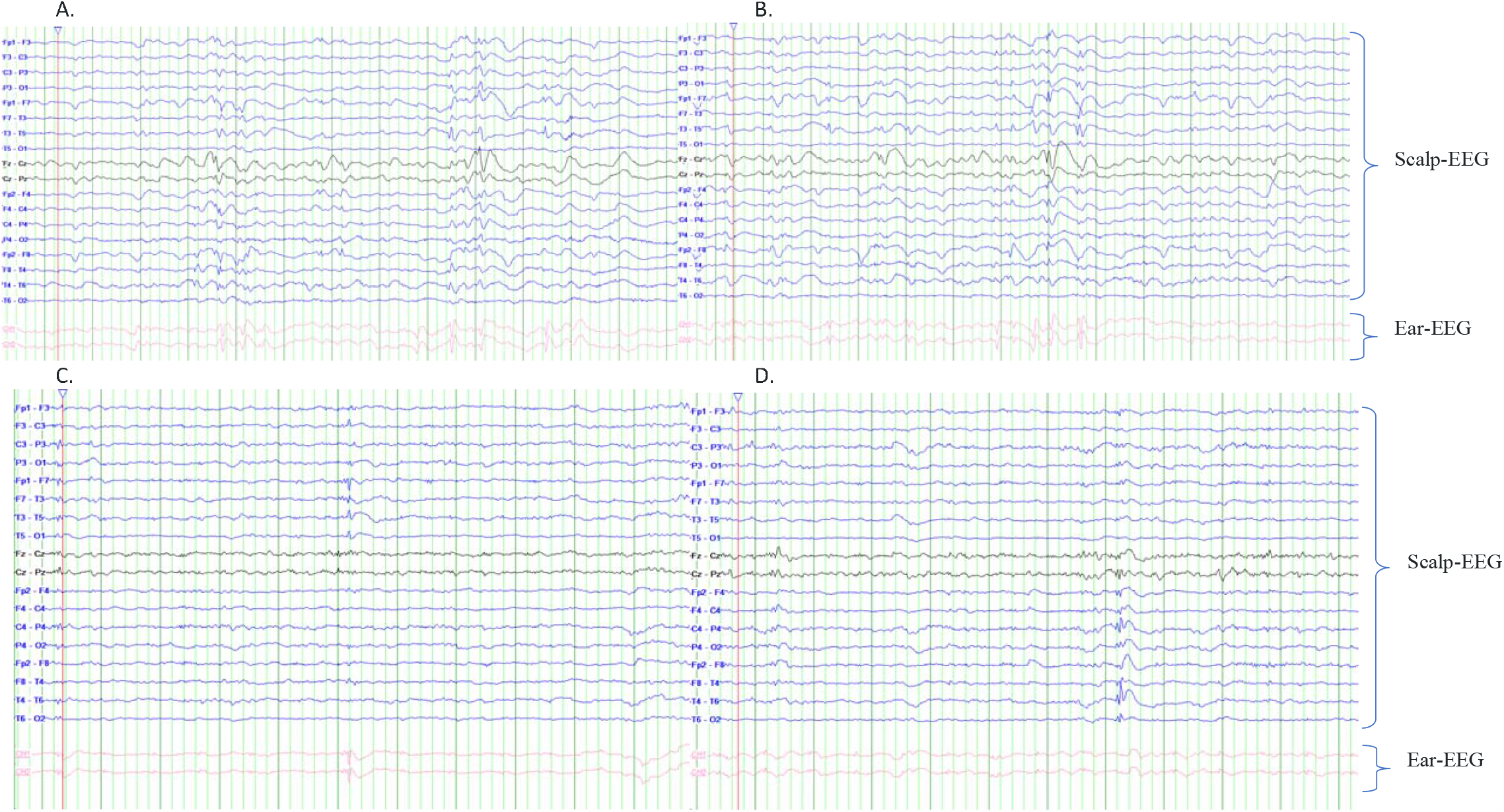
Comparison of the full montage of scalp EEG and ear EEG. (A and B) shows the EEG study of a patient with slow spike-wave complex discharges in the right frontotemporal region. (C and D) the study of another patient with independent interictal discharges in anterior temporal regions, the patient was wearing the device on the right side. The scalp channels are shown in the common bipolar montage synchronised at Natus Neuroworks. The two channels of the earEEG device are shown in pink.

### 2.4. EEG data processing

The EEG data processing in both v-EEG and ear-EEG consisted of different steps:

⍰ Alignment and synchronization of timeline for both EEG data. EEG data was recorded in two different systems, so it was needed to align the onset of the intervals. The difference in sampling rate was determined and adjusted to a common resampling.
⍰ Discarding of intervals with artefacts. Raw data obtained from video-EEG recordings contains noise and electrical interferences, which distort the recorded signals. Artifacts in EEG recordings are disturbances in brain signal, which are outlier’s measurement not generated within the brain. External artifacts are caused frequently from aberrant technology such as electromagnetic interference and disconnection of the electrode box (for instance, displacement in the skin-electrode contact). Internal artifacts may occur due to changes of the potential between electrodes as consequences from eye movement or muscular activity. We implemented a set of filters to remove the electrical power line interference at 50 Hz and the baseline drift at 0.5 Hz detecting only the frequencies that are from brain activity. Furthermore, EEG experts performed visual detection of this artifacts, and these were automatically detected and rejected. Several methods have been described for the automation of this process. We use the amplitude thresholding method, calculated as three to five times the standard deviation of the signal.
⍰ Feature extraction from individual artifact-free time series of the EEG recordings. The signals were transformed into statistical descriptive values, named features. This data is segmented into fixed sized windows, which have a certain degree of overlap between contiguous segments. Each of the windows is labelled using a binary method, interictal segments are associated to a zero value and preictal segments are labelled with a unitary value. The windows are 60 seconds length with a 50% overlap. In total, over 150 features have been implemented in the algorithm for different subbands. The implemented features can be divided in the following main groups:
  - Energies: They measure the degree of electrical activity for a given range of frequencies. Such values are scared to different magnitudes to exploit the non-linear properties of the energies.
  - Complexity measurements: describing the variability of the signal, finding trends and calculating the degree of predictability of such trends.
  - Centrality measurements: Describing the values distribution of the signal.
  - Connectivity measurements: Calculating the degree of synchronization between the two channels, analyzing the similarity of the patterns.

#### 2.4.1 EEG features

The power, energy, entropy, and kurtosis of an electric signal play a crucial role in algorithm generation for various applications in electrical engineering and signal processing. These signal characteristics provide important information that can be utilized in the design, analysis, and optimization of algorithms. Let’s explore the relevance of each of these measures in the algorithm generation:

- Power: The power of an electric signal determines its intensity or strength. In algorithm generation, power is often used for signal normalization, scaling, or dynamic range adjustment. It helps in ensuring that the signal is within a suitable range for subsequent processing steps. Power-based algorithms can be designed to detect changes in signal power, identify peaks or transients, or even classify signals based on their power characteristics.
- Energy: Energy represents the total amount of work done or transferred by a signal over a given period. In algorithm generation, energy is often utilized for feature extraction, event detection, or activity recognition. Algorithms can be designed to analyze the energy distribution within a signal, identify energy peaks or bursts, or to calculate the total energy of a signal segment. Energy-based algorithms are commonly used in applications such as speech recognition, vibration analysis, and power system monitoring.
- Entropy: Entropy measures the amount of uncertainty or randomness in a signal. In algorithm generation, entropy is widely used for feature extraction, data compression, or anomaly detection. Algorithms can be designed to analyze the entropy of a signal to identify regular patterns, to detect sudden changes or abnormalities, or to classify signals. Entropy-based algorithms find applications in fields such as image processing, data mining, and pattern recognition.
- Kurtosis: Kurtosis describes the shape or distribution of a signal. In algorithm generation, kurtosis is valuable for feature extraction, outlier detection, or classification tasks. Algorithms can utilize kurtosis to identify signals with heavy-tailed distributions, detect anomalous events or outliers, or differentiate between different signal classes based on their kurtosis values. Kurtosis-based algorithms are applied in areas such as financial analysis, fault diagnosis, and environmental monitoring.

By incorporating power, energy, entropy and kurtosis with other features such as delta band average power, spectral centroid, zero crossing, cross-correlation, mutual information, geometric mean, harmonic mean and Hurst fractal dimension into algorithm generation, engineers and researchers can develop algorithms tailored to the specific characteristics of electrical signals. These measurements provide information about the intensity, randomness, distribution and outliers present in the signals, allowing a more accurate and efficient algorithmic processing.

### 2.5. Visual correlation

The present study received authorization from an accredited Ethics Committee for Medicines Research (CEIm); more specifically, the Regional Ethics Committee for Medicines Research of the Community of Madrid. The period of conservation of the data subject to analysis will be limited to the legal periods established for this purpose by the regulations on data protection and clinical research. Subsequently, said information will be kept in a blocked form without the possibility of further processing, unless it is reused under the terms and conditions permitted by the applicable ethical criteria, by the applicable legislation on research with clinical documentation (among others, Law 41/2002, of 14 November, basic law regulating patient autonomy and rights and obligations regarding clinical information and documentation) and by the regulations on the protection of personal data (among others, Organic Law 3/2018, on Data Protection and Guarantee of Digital Rights).

### 2.6. Statistical analysis

For each patient, a random sample of time frame was selected, and the correlation between scalp and ear-EEG was calculated, according to a custom Matlab script, using a sliding window technique to calculate the power of the scalp-EEG and ear-EEG signals. The time frame sample was of 2 minutes in the experiments which last below 15 minutes and of 5 minutes in the experiments beyond 30 minutes. A Matlab correlation coefficients function (corrcoef) was used to make the final correlation. The ‘corrcoef’ function measures the linear dependence between the two signals, using the Pearson correlation coefficient.

From each patient, several random samples were obtained and labelled by experts who determined in patients if there was seizure activity. In controls, time frames were labelled as repose, minimum 15 minutes with relaxing music, sleeping, minimum 60 minutes, and mental activity, minimum 15 minutes of working with Sudoku or similar. To analyze the data, several different sensitivity analyses were performed employing different subsets of patients and registers: 1) Whole signal analysis, which included all the registers (subject results, SR), 2) Filtered results (FR) which discarded the segments with visible artifacts in the scalp or ear EEG signal.

Mixed linear models were employed to analyze the collected data. Thus, it was possible to capture variability within patients (intra-subject variability) as well as between patients (inter-subject variability). For the estimation of main correlation, a linear mixed model was obtained using the filtered correlation as the predicted variables and the ID of the patients as random variable. To study the differences between patients and controls we added the group as fixed effect. Finally, to detect differences in correlation between types of activities, we fitted a linear mixed model employing the activity as fixed variable.

Categorical variables are expressed as the total number (N) and percentage. For the estimation of correlations, the mean estimate of the difference and the 95% confidence interval are shown. Alpha was set as 0.05. All analyzes were performed in R version 4.0.2 using the lme4 and lmerTest packages.

## 3. RESULTS

Thirty subjects (56.7% women) from 20 up to 62 years fulfilled the inclusion criteria. Sixteen had drug-resistant focal epilepsy, and the remaining were controls. Two individuals in the epilepsy group were excluded from the analysis due to the lack of clinical data (final total n=28, 14 patients, 14 controls). Out of the 14 patients, the location of epilepsy was bilateral frontal (n=7), anterior temporal (n=5) and posterior temporal (n=2) (table 1). Fifteen seizures were detected by v-EEG and 14 seizures were registered with both systems simultaneously. In total 545 hours of v-EEG and 387 hours of ear EEG were recorded.

**Table 1:** Patient characteristics.

| Age | Sex | Side | ASM(s) | ASM(s) previous | EEG previous | Seizure frequency | Seizure semiology | v-EEG duration (h) | Ear-EEG duration (%) | Seizure recorded (Ear/total) |
| --- | --- | --- | --- | --- | --- | --- | --- | --- | --- | --- |
| 21-25 | M | R | LEV, LCM | N/A | Right anterior temporal (T4 > F8) spike-wave complexes | 3-4/weekly | Epigastric aura, unresponsiveness, automotor behaviors and later axial tonic posture | 44 | 74% | 0/0 |
| 26-30 | M | R | LCM, PER | ESL, LTG, BRV | Right anterior temporal (T4 > F8) spike-wave complexes | 5-6/monthly | Epigastric aura, unresponsiveness and automotor behaviors | 46 | 78% | 1/2 |
| 56-60 | F | L | VPA, CNB | CBZ, LCM, PER | Bilateral frontal 2-3 Hz spike-wave complexes left predominant. | 1-2/daily | Tonic and hyperkinetic seizures | 60 | 58% | 0/0 |
| 61-65 | M | R | PER, LTG | PHT, CBZ, LCM | Bilateral frontal 3-4 Hz spike-wave complexes left predominant. | 6/monthly | Axial tonic posture and upper limbs in flexion and late unresponsiveness | 88 | 72% | 1/1 |
| 46-50 | M | L | LEV, LTG, VGB, VPA, PB | CBZ, PER | Bilateral frontal 3-4 Hz spike-wave complexes left predominant. | 4-5/daily | Atonic and tonic seizures with multiple falls, atypical absences | 54 | 69% | 2/2 |
| 26-30 | F | L | CNB, BRV, LCM | CBZ, PER, LEV, LTG | Right anterior temporal (T4 > F8) spike-wave complexes | 2/monthly | Perceptual disturbance, depersonalisation, jamais-vu, unresponsiveness and prolonged post-critical | 48 | 45% | 1/1 |
| 41-45 | F | L | CBZ, BRV, CLB | LTG, LEV, PER | Bilateral frontal 2-3 Hz spike-wave complexes left predominant. | 3-4/monthly | Tonic-clonic bilateral | 43 | 100% | 0/0 |
| 21-25 | M | L | CLB | LCM, ESL, CBZ, LTG | Right anterior temporal (F8>T6>T4) spike-wave complexes | 4-6/weekly | Change in facial mimics, pallor, disconnection from the environment and bilateral upper limbs exploratory automatisms. | 41 | 61% | 3/3 |
| 31-35 | F | R | LTG, VPA, CNB | LCM, ESL, BRV, CBZ, PER, LEV | Right post-temporooccipital spike-wave complexes | 5-7/monthly | Fear reaction, unresponsiveness and tonic posture axial and upper limbs | 43 | 60% | 0/0 |
| 16-20 | F | L | ESL, PER, CNB | VPA, OXC, TPM, LCM | Left post-temporooccipital intermittent sharpes | 2-3/monthly | Tonic and hyperkinetic seizures | 12 | 100% | 3/3 |
| 61-65 | M | R | LEV, LCM, PER | ESL, BRV, TPM | Intercritical epileptiform abnormalities in both temporal regions | 10/monthly | Unresponsiveness, tonic posture left upper limb, post-ictal aphasia. | 10 | 100% | 0/0 |
| 21-25 | F | R | VPA, LEV,<br>LCM | CBZ, CLB, ETS,<br>RUF, VPA,<br>LTG, TPM,<br>LEV. | Bilateral frontal 2-3 Hz spike-wave<br>complexes left predominant. | 5-6/weekly | Unresponsiveness, tonic posture<br>and elevation both upper limbs | 12 | 75% | 1/1 |
| 21-25 | M | L | VPA, LCM,<br>LTG | CBZ, CLB, LEV,<br>PER | Bilateral frontal 2-3 Hz spike-wave<br>complexes right predominant | 2-3/daily | Unresponsiveness and head drop | 20 | 80% | 1/1 |
| 21-25 | F | L | ESL, PER,<br>LCM | VPA, CBZ, LTG | Bilateral frontal 2-3 Hz spike-wave<br>complexes right predominant | 1-2/daily | Unresponsiveness, tonic posture<br>axial and upper limbs | 24 | 86% | 1/1 |

Abbreviations. ASM: Anti-seizure medication, BRV: Brivaracetam, CBZ: Carbamazepine. CNB: cenobamate, CLB: Clobazam, CLZ: Clonazepam, EEG: electroencephalogram, ETS: Ethosuximide, ESL: Eslicarbazepine, LCS: Lacosamide, LEV: Levetiracetam. LTG: Lamotrigine. OXC: Oxcarbamazepine. PHT: Phenytoin, PER: Perampanel, RFN: Rufinamide, TPM: Topiramate VPA: Valproate, ASM: Antiseizure medication, EEG: electroencephalogram, v-EEG: scalp video EEG, ear-EEG, M: male, F: female, R: right, L: left, N/A: not available.

AC in the whole sample was 0.88 [0.86 - 0.90], and it was 0.90 [0.88 - 0.91] in the filtered results. Epileptic patients had an AC of 0.88 [0.87 - 0.91] in filtered results and 0.87 [0.83 - 0.89] in SR, while in controls AC was 0.90 [0.88 - 0.92] in filtered results and 0.89 (CI95% 0.87 - 0.92) in SR. No differences in the correlation of signals were found between controls and patients in the FR (-0.01 [- 0.04;0.01], p=0.261) or the SR (-0.03 [-0.06;0.01], p=0.09).

In patients with epilepsy, no differences in AC were noted between ictal (AC 0.88 [0.83 – 0.92]) and interictal periods (AC 0.89 [0.87 – 0.91], mean difference in AC -0.02 [-0.06; 0.02], p=0.352) (fig. 4). However, AC in controls was found to be smaller during sleep (SR 0.88, FR 0.88) compared to repose (mean difference in AC -0.04 [-0.08; -0.01], p=0.01). No artifacts were detected in the studied timeframes of sleep.

**Figure 4:**
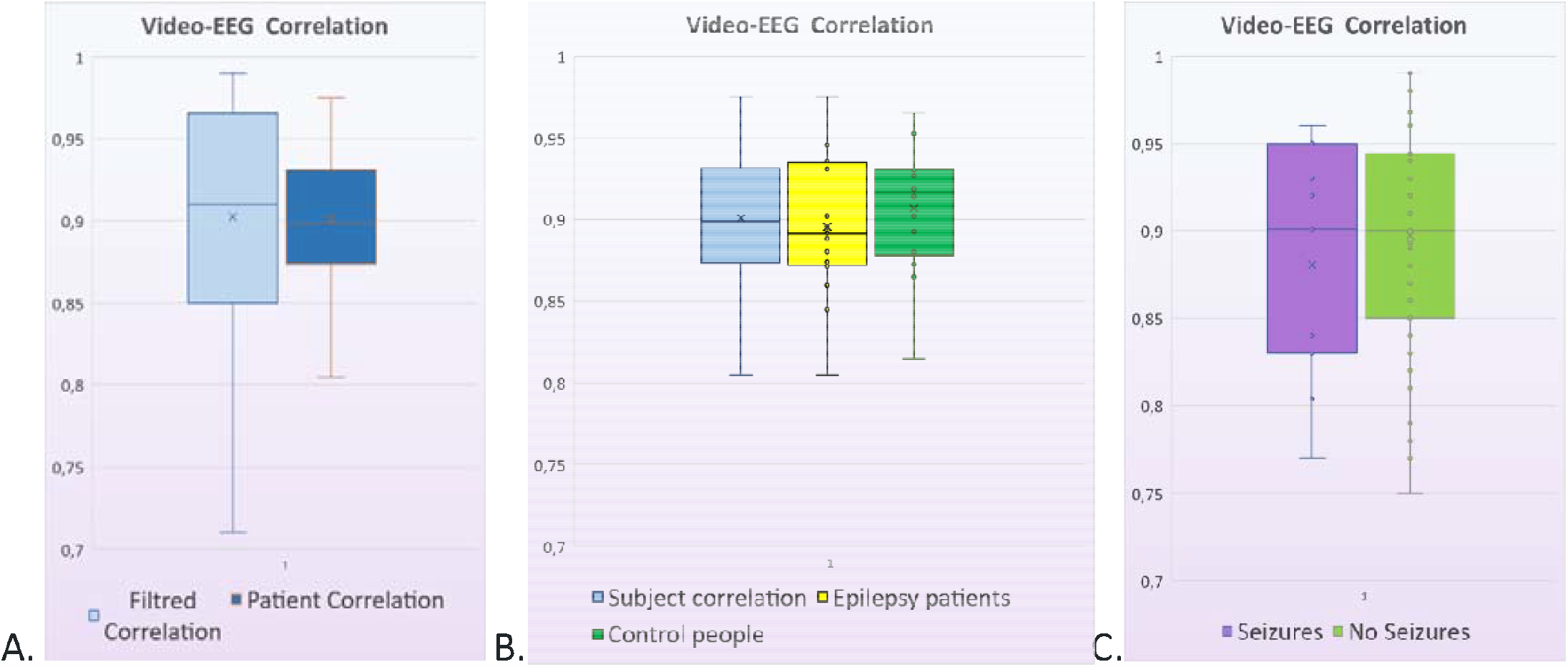
(A) Correlation of energies of 0.90 in filtered results (FR) and 0.88 in subject results (SR). (B) In the epilepsy group, an AC of 0.88 in FR and 0.87 in SR, and the control group 0.90% in FR and 0.89 in SR. (C) Results distributed by ictal periods with AC of 0.88 and interictal periods of 0.89.

Visual comparison of raw (fig 5A) signal, spikes and waves (fig 5B), as well as time evolution of features spectrograms (fig 5C) and distribution of features (fig 5D), showed the signal between v-EEG and ear-EEG was similar.

**Figure 5:**
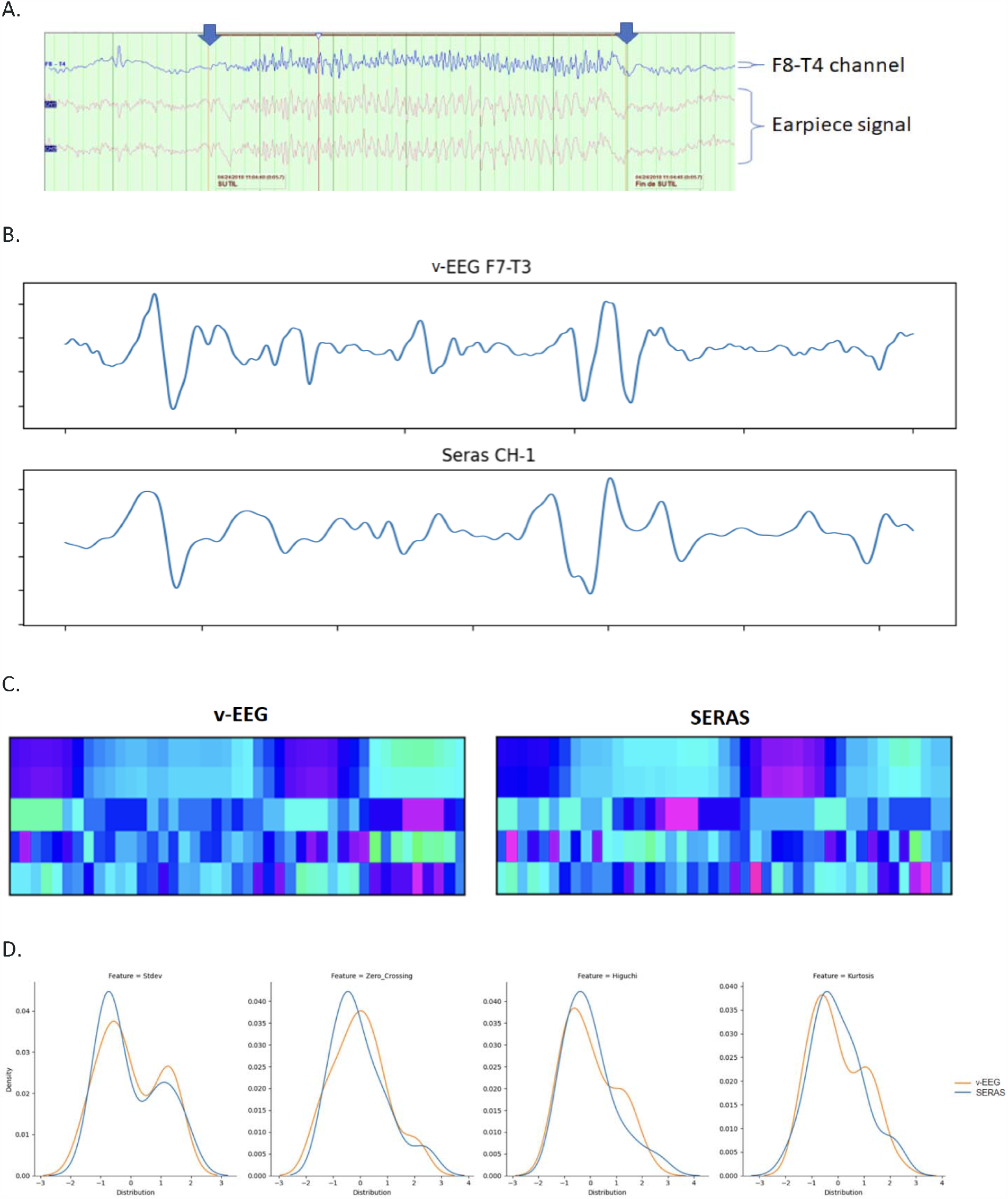
Visual comparison between temporal anterior scalp-EEG and ear-EEG in a conventional viewer (A), with the temporary viewer for ear-EEG (B), between spectrograms generated from the signal of both (C) and after analysis of some features (D). Panel C represents the temporal evolution of five extracted features (Y axis) over time (X axis). Note that both spectrograms present a high overlapping of the colours.

No adverse effects of the device have been reported. Most patients have tolerated the device well, with only four cases mentioning a sensation of presence in the ear, which was well tolerated and disappeared within a few hours.

## 4. Discussion

Our study shows how our ear EEG device can be used to detect the brain electrical activity. The findings that support our claim are based on the high energy correlation between scalp EEG and our device in different groups of individuals (healthy subjects and epileptic patients) and contexts (ictal vs interictal, relax vs activity).

To our known, our device is one of the first to demonstrate such high correlation with scalp EEG (0.90 [0.88 - 0.91] in filtered results). Previous studies demonstrated the feasibility of ear EEG to identify energy patterns linked to focal seizures, but with a much lower energy correlation between signals (0.54, CI95 0.51–0.57)(10). Although the size of the correlation itself does not demonstrate if the information to detect the seizure is retained in the ear-EEG, previous studies showed seizure onset could be detected from inspection of the ear-EEG recording in a similar proportion of cases to scalp EEG, demonstrating that ear EEG keeps the necessary information (10) The differences in the correlation with prior studies may be attributed to the differences in the position of the reference electrodes (our study inside de ear channel, in prior studies with one on the pinna) and the different methods employed to calculate the correlation (our study with the energy, in prior studies (10) with the raw time-frequency signal).

In this study, we also investigated the capability of our device to detect the scalp-EEG information in different contexts such as ictal and interictal periods, sleep and wakefulness. We found energy correlation was similar in all the different situations and we only found a lower correlation during sleep was than in repose (-0.04 [-0.08; -0.01], p=0.01), in FR. Further studies are needed to confirm the results. We were unable to demonstrate differences in energy correlations between other contexts, showing a high stability in the signal despite different states. This is in line with prior studies, which demonstrated how the ear EEG was capable of detecting events accurately (12)

There are some differences between our device and previously reported devices in the literature which may also make the comparison of our findings. First, our device only employed 3 sensors, compared to other EEG configurations which employ up to 5 sensors (9). Our device has 2 channels of EEG data, with only one reference for both channels, which makes it easier to industrialize but more sensible to artifacts. The position of the sensors is also important. Unlike other devices, mjn-SERAS does not have any sensor placed on the ear pavilion, having all sensors inside the ear canal. This configuration provides a shorter distance with the focus but with a lower voltage. The most efficient configuration of ear EEG is still to be defined, as there are no studies comparing different sensors and positions.

Despite the strengths of our study, there are some limitations. First, the limited number of participants in the study may restrict the generalizability of the findings. Also, the recordings were obtained in a controlled environment, which may not fully reflect real-life conditions. However, the comparison with a gold standard in an experienced epilepsy monitoring unit, improves the reliability of the device’s performance to record EEG data for epilepsy patients.

SERAS-EEG study provides technical support for use of the mjn-SERAS to record EEG signals compared to the gold standard.

The high degree of similarity of the recordings compared suggests that the use of mjn-SERAS may be suitable for ambulatory EEG functions, seizure detection or prediction, but due to the use of only two recording channels it does not allow for epilepsy diagnostic utility. The prolonged use of these registers will increase the possibilities of collecting unusual, uncommon, or difficult to classify events, allowing the study of their characteristics and comparison with the usual events. Assisted by a calendar and manual seizure registers, it allows the patient to have essential information to pass on to his or her physician at subsequent visits. Furthermore, a large amount of data is generated which allows the use of processing algorithms and the growth of epilepsy and seizure databases, for the continuous improvement of processes and algorithms that work with this data, both in real time for seizure detection and prediction and in scientific research.

A prospective, multicenter, pilot clinical trial is currently in progress to evaluate the mjn-SERAS in real life to anticipate seizures and describe improvements in different areas of personal development of epilepsy patients. This study proposes the use of the mjn-SERAS device during the day-to-day life of the patient to analyze its performance in generating alerts in the case of the possibility of a high risk of epileptic seizures and to evaluate the concordance and prediction of the generation of early detection seizure alarms, prior to the identification of these clinically manifested events and collected by the patient or their relatives. In addition to the evaluation of these preventive alerts that we can associate with the physical sphere of the concept of health, we are going to determine, study and analyze the impact of the mjn-SERAS device on psychic and/or mental well-being as well as its repercussion on the social well-being of people with refractory epilepsy.

## Data Availability

All data produced in the present study are available upon reasonable request to the authors

## Author Contributions

G. T-G wrote the first draft of the manuscript and evaluated the patients. D.B. conceived the original idea and revised the manuscript. A.V performed the statistical analysis and revised the manuscript. X. R, J.V., L.M and J.L performed the signal analysis and revised the manuscript. A.T and A. A-S revised the manuscript.

